# Vitamin D Status & Latitude Predict Brain Lesions in Adrenoleukodystrophy

**DOI:** 10.1101/2021.12.26.21268422

**Authors:** Keith van Haren, Jacob Wilkes, Ann B. Moser, Gerald V. Raymond, Troy Richardson, Patrick Aubourg, Timothy W. Collins, Ellen M Mowry, Joshua L. Bonkowsky

## Abstract

A subset of boys with X-linked adrenoleukodystrophy (ALD) develop inflammatory demyelinating brain lesions. Risk factors are largely undefined. We used two independent cohorts to assess whether low vitamin D status predicts lesion development. In our first cohort, we measured 25-hydroxyvitamin D in 53 plasma samples from 20 pre-lesional ALD boys followed at two centers; half subsequently developed lesions. In our second cohort, we measured latitude (using home ZIP code) among 230 ALD boys in a database of 51 US pediatric hospitals; over half developed lesions. In regression models, low plasma vitamin D and northerly latitudes independently predicted ALD brain lesions.

## INTRODUCTION

X-linked adrenoleukodystrophy (ALD) is caused by mutations in a single gene (*ABCD1*) which encodes a peroxisomal fatty acid co-transporter, leading to accumulation of very long chain fatty acids in adrenal cortex, blood, and brain tissue.^1^ Thirty to forty percent of boys with ALD will develop a disease state termed cerebral ALD (cALD); incidence is concentrated in the first decade.^2,3^ cALD is marked by progressive inflammatory demyelinating brain lesions. Left untreated cALD culminates in a vegetative state and death within a few years of onset.^4^ Small, early-stage brain lesions can be halted with hematopoietic stem cell transplant.^4,5^ Although the presence of a pathogenic *ABCD1* variant confirms an ALD diagnosis, the risk for cALD is discordant in siblings or even identical twins.^1^ This suggests a role for environmental or epigenetic risk factors. Understanding etiology would be valuable for surveillance, treatment, and prevention.

Notably, ALD brain lesions share key similarities with multiple sclerosis lesions, including a predilection for the corpus callosum and an inflammatory demyelinating histology characterized by a leading edge of activated microglia, lipid-laden macrophages, and trailing lymphocytes.^6^ We reasoned that shared histology could suggest shared risk factors. Lower levels of vitamin D exposure have been robustly linked to higher risk of multiple sclerosis diagnosis, subsequent brain lesions, and improved prognosis.^7-11^ Trials of vitamin D supplementation in multiple sclerosis have demonstrated modest benefits, including a reduction in the appearance of new brain lesions.^12,13^ Building on this premise, we investigated whether two established proxies for vitamin D exposure, plasma 25-hydroxyvitamin D status and north-south latitude, were correlated with the risk of developing cALD brain lesions.

## METHODS

The Institutional Review Boards (IRB) of Stanford University, Kennedy Krieger Institute, Institut National de la Santé et de la Recherche Médicale (INSERM), and the University of Utah, exempted this study as non-human research.

To assess correlation of pre-morbid plasma 25-hydroxyvitamin D and subsequent development of cALD, we analyzed previously biobanked plasma samples collected as part of longitudinal clinical cohort studies of ALD boys at two centers (Kennedy Krieger Institute, Baltimore, Maryland, USA & INSERM, Paris, France) between May 1, 1999 and November 8, 2012. All patients were followed with serial MRIs every 6-12 months to screen for brain lesions according to standard care guidelines.^1^ ALD brain lesions were defined as any gadolinium-enhancing white matter lesion on brain MRI. Plasma samples were collected longitudinally and stored at -80°C. Inclusion criteria for plasma analysis were: (i) molecular/biochemical diagnosis of ALD, (ii) male sex, (iii) age <11 years at sample collection (corresponding with period of maximal risk for cALD^2,3^), (iv) normal brain MRI prior to plasma collection, and (v) availability of plasma samples from at least two timepoints prior to study endpoint. Study endpoints were (i) identification of cALD brain lesion on MRI or (ii) no ALD brain lesion identified at time of last follow-up. We excluded samples collected after a brain lesion was identified. We measured 25-hydroxyvitamin D levels (Heartland Assays, Inc.) and C26:0 lipid levels (Kennedy Krieger Institute) using tandem mass spectrometry. Lipid levels were measured at the time of plasma collection. We used a two-tailed Mann-Whitney U-test to assess differences between groups (Prism Graphpad v8.4). We used a logistic regression analysis to estimate the odds of developing brain lesion based on each patient’s average 25-hydroxyvitamin D level (STATA v11).

To assess correlation of latitude and cALD, we determined the total number of ALD boys in the Pediatric Health Information System (PHIS) database. Patients had to be male, age <19 years, with a valid US ZIP code, and presenting between October 1, 2015 and June 30, 2019; with an ICD10 code for ALD, including E71520 - Childhood cerebral X-linked adrenoleukodystrophy; E71521 - Adolescent X-linked adrenoleukodystrophy; E71522 – Adrenomyeloneuropathy; E71528 - X-linked adrenoleukodystrophy NEC; and E71529 - X-linked adrenoleukodystrophy NOS. We assessed for the primary clinical outcome, the development of brain lesions, by the presence of an ICD9 or ICD10 code for at least one of the following categories indicating CNS involvement: bone marrow transplant/hematopoietic stem cell transplant; cerebral degeneration or neurological dysfunction; or cortical visual loss/blindness (full list in ***Supplemental Table 1***). We used ZIP code to determine latitude; we collapsed latitude into 2° ranges to allow a minimum of two ALD cases in each category. We excluded latitude ranges that had no ALD cases (19-24° north and >49° north). For each latitude range, we calculated the unadjusted probability of cALD among total ALD cases. We used a logistic regression model to calculate the probability of cALD among ALD patients adjusting for the size of PHIS population. Finally, we modeled the relationship between latitude ranges and probability of cALD compared to ALD using logistic regression, adjusting for total PHIS population size in that latitude (SAS v9.4).

## RESULTS

We identified 20 ALD boys with plasma samples collected at 53 timepoints over an average of 3.8 ±3.1 years between first plasma collection and clinical endpoint. Ten boys developed brain lesions during observation (median Loes score 1.0). Although plasma samples were collected at similar ages in both groups, patients who never developed cALD were followed longer (***Table 1***). Average 25-hydroxyvitamin D levels were significantly lower among boys who developed cALD than those who did not (median 28.9 vs 36.6 ng/ml); *p*=0.019 (***Table 1***). For each 10ng/mL decrease in 25-hydroxyvitamin D level, the odds ratio for developing cALD was 6.94 (95% confidence interval, 1.05, 45.8); *p*=0.044. Fatty acid biomarkers associated with ALD were available for 16 patients; they were similar across groups (***Table 1***).

**Table 1.**
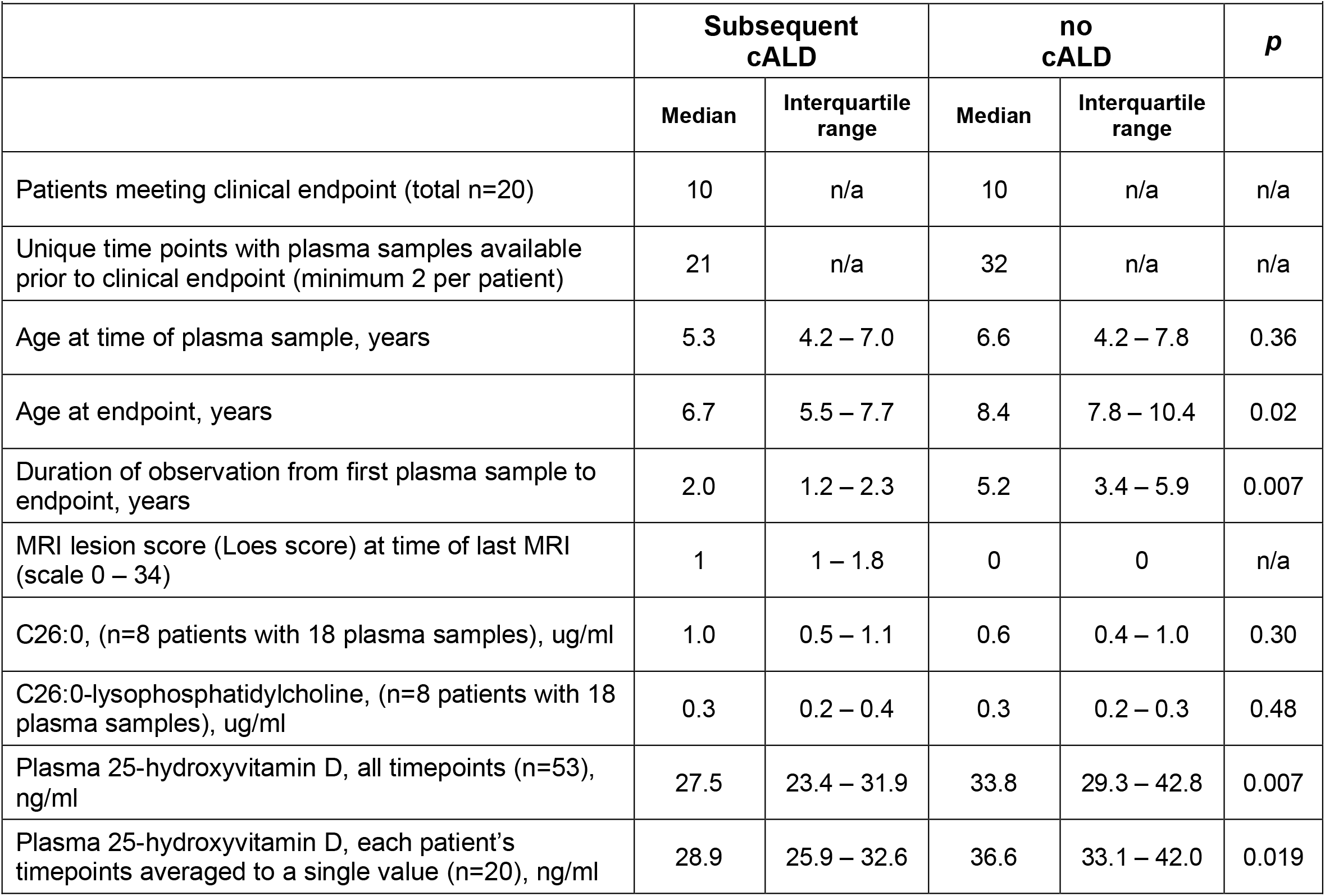
Demographics and plasma levels from 20 ALD boys at two academic medical centers. All boys had normal brain MRIs at the start of observation and were followed with serial MRIs to detect early-stage cALD. Clinical endpoints assigned according to whether patient developed cALD during period of observation.

For latitude analysis, we identified 230 ALD boys across 28 states and 21º of latitude; 57% of boys (n=132) developed cALD. Each 2º increase in latitude conferred an odds ratio of 1.17 (95% confidence interval, 1.01, 1.35); *p*=0.036 for developing cALD (***Table 2; Figure 1***).

**Table 2:** Incidence risk of cALD and latitude. cALD rates at each latitude range are shown before and after adjustments for Pediatric Healthy Information System (PHIS) population. The inclusion of latitude in the regression model significantly affected cALD rates. To protect patient confidentiality, we have not provided numerators (cALD patients) and denominators (all ALD patients) by latitude category since almost half the latitude ranges have either (i) fewer than 10 patients or (ii) fewer than 3 hospitals.

| Degrees Latitude* | Total PHIS Population | Unadjusted probability of cALD (% with cALD) | Adjusted** probability of cALD – latitude excluded from model (% with cALD) | Adjusted** probability of cALD – latitude included in model (% with cALD) |
| --- | --- | --- | --- | --- |
| 25°-26° | 101,841 | 50.0% | 54.7% | 36.7% |
| 27°-28° | 106,949 | 25.0% | 54.8% | 40.2% |
| 29°-30° | 323,873 | 30.0% | 55.4% | 42.8% |
| 31°-32° | 438,066 | 50.0% | 55.5% | 46.4% |
| 33°-34° | 881,280 | 40.0% | 56.6% | 48.2% |
| 35°-36° | 472,796 | 43.8% | 55.6% | 53.9% |
| 37°-38° | 437,657 | 75.8% | 55.5% | 58.0% |
| 39°-40° | 1,060,064 | 56.0% | 57.2% | 58.5% |
| 41°-42° | 698,861 | 72.1% | 56.1% | 64.2% |
| 43°-44° | 156,815 | 50.0% | 54.8% | 69.9% |
| 45°-46° | 51,321 | 0.0% | 54.6% | 73.4% |
| 47°-48° | 72,553 | 60.0% | 54.6% | 76.3% |

\*Collapsed to get at least 2 cases of ALD in each category
\*\*Adjusted for PHIS population

**Figure 1.**
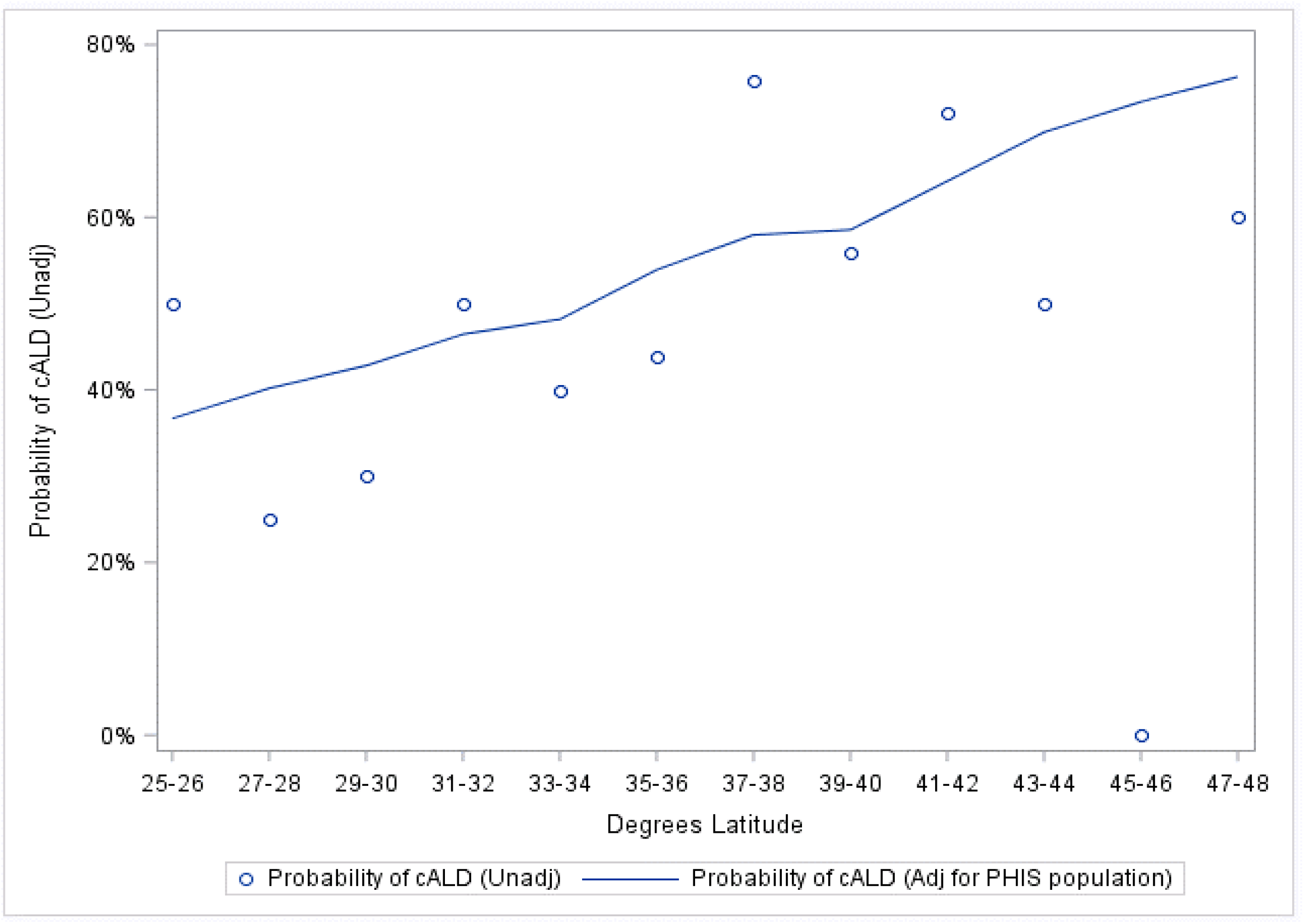
Graph of incidence risk of an ALD brain lesion compared to latitude. Each 2º increase in latitude conferred an odds ratio of 1.17 (95% confidence interval 1.01, 1.35); *p*=0.036 for developing cALD.

## DISCUSSION

Using independent cohorts, we found that ALD boys with lower pre-morbid plasma levels of 25-hydroxyvitamin D, or more northerly latitude of residence, were more likely to develop inflammatory brain lesions (cALD). These findings offer complementary lines of evidence implicating vitamin D and/or ultraviolet light exposure as a regulator of cALD risk.

We derived the concept of vitamin D as risk factor for cALD based on histologic similarities between ALD and multiple sclerosis brain lesions.^6,14^ In multiple sclerosis, vitamin D’s therapeutic mechanism is attributed to its role in immune homeostasis.^12,14^ Similar immunologic mechanisms are plausible in reducing risk of developing the inflammatory demyelination that similarly characterizes cALD. However, because the ALD genotype disrupts peroxisomal fatty acid metabolism and causes elevated very long chain fatty acid levels, our findings may implicate vitamin D in mediating fatty acid metabolism and/or its role in immunologic homeostasis.^15-17^

Limitations of our study include the retrospective nature of our analyses and modest sample sizes, the latter of which limits our ability to assess for putative confounders. Although these analyses represent, to our knowledge, the first two assessments of vitamin D as a risk factor for cALD, prospective studies with or without intervention would further validate and refine the magnitude of risk associated with low vitamin D status in boys with an ALD genotype.

The advent of universal newborn screening for ALD has increased the number of ALD boys who can benefit from prospective MRI surveillance. Because ALD brain lesions rarely manifest before 2 years of age, widespread newborn screening could facilitate a trial of early life vitamin D supplementation to test its potential as a preventive therapy against ALD brain lesions.

In summary, we describe two complementary lines of evidence implicating low vitamin D status as a risk factor for the development of brain lesions among ALD boys. Our findings also support the possibility of shared disease mechanisms in cALD and multiple sclerosis.

## Data Availability

All data produced in the present study are available upon reasonable request to the authors

## ACKNOWLEDGEMENTS

JLB was supported by NIH grant 3UL1TR002538, and by the Bray Presidential Chair in Child Neurology research.

KV was supported by Child Neurology Society Scientific Award; NIH grant K23NS087151; the Tashia and John Morgridge Endowed Faculty Scholarship in Pediatric Translational Medicine of the Stanford Maternal & Child Health Research Institute; gifts to the Lucile Packard Foundation, the Lenail-Yoler Family, the Adler Family, the Senkut Family, and from Arrivederci ALD.

## AUTHOR CONTRIBUTIONS

KV and JLB conceptualized and designed the study and drafted the manuscript.

JLB, EMM, KV, TWC, JW analyzed the data and edited the manuscript for intellectual content.

AM, GVR, and PA supervised and curated the longitudinal cohorts including specimen management, specimen selection, and associated clinical data.

---

**Supplemental Table 1.**
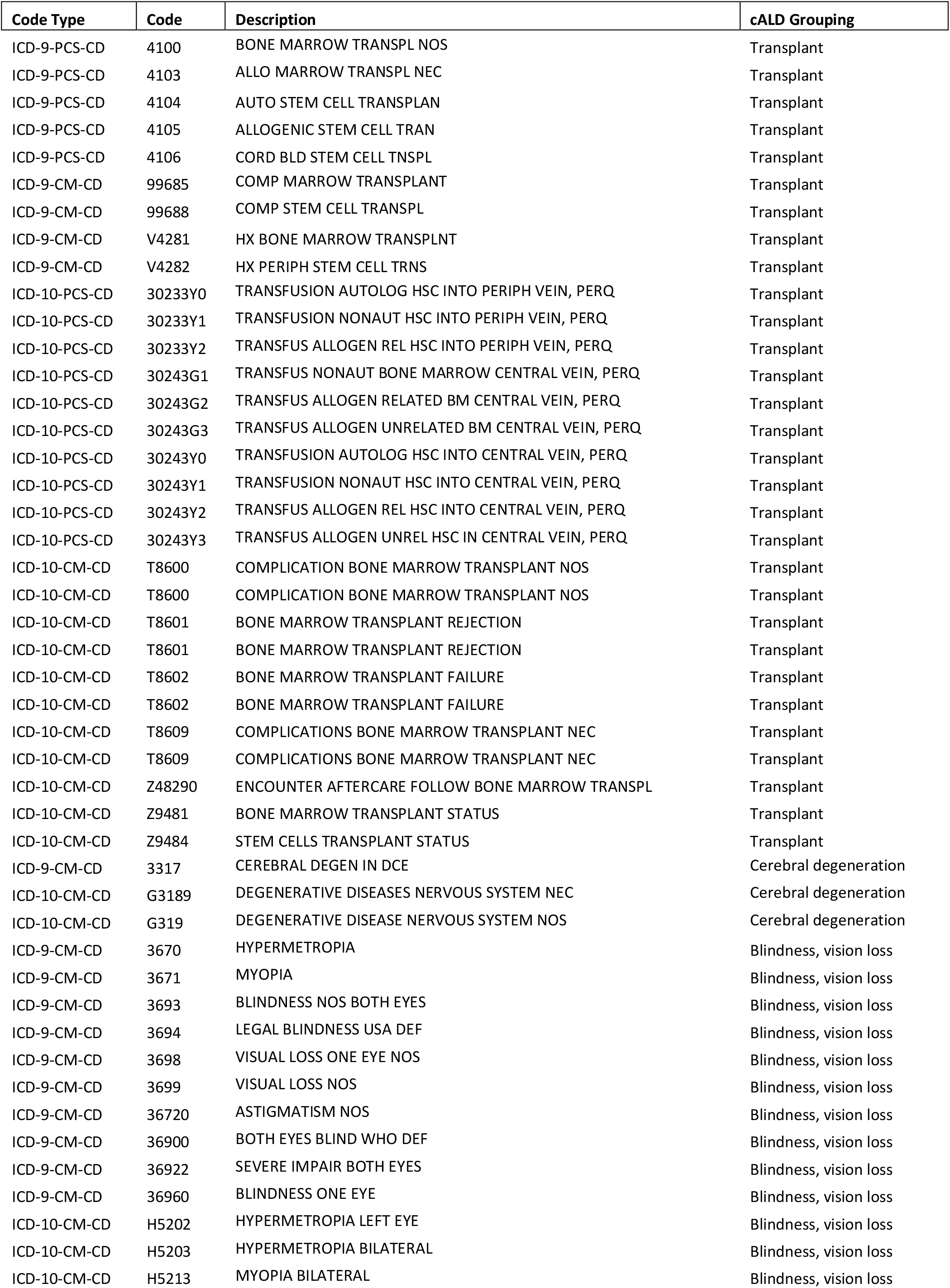
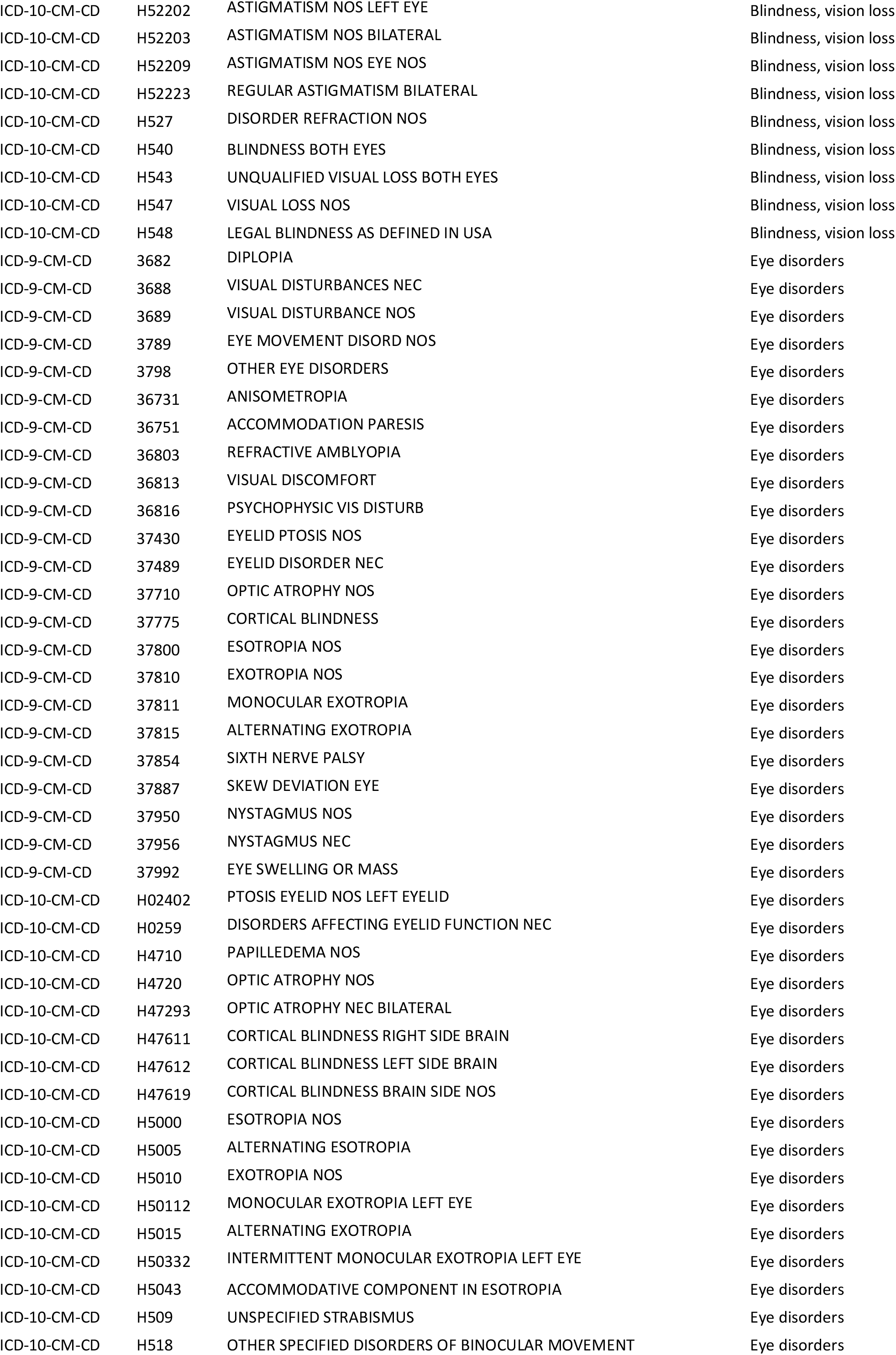
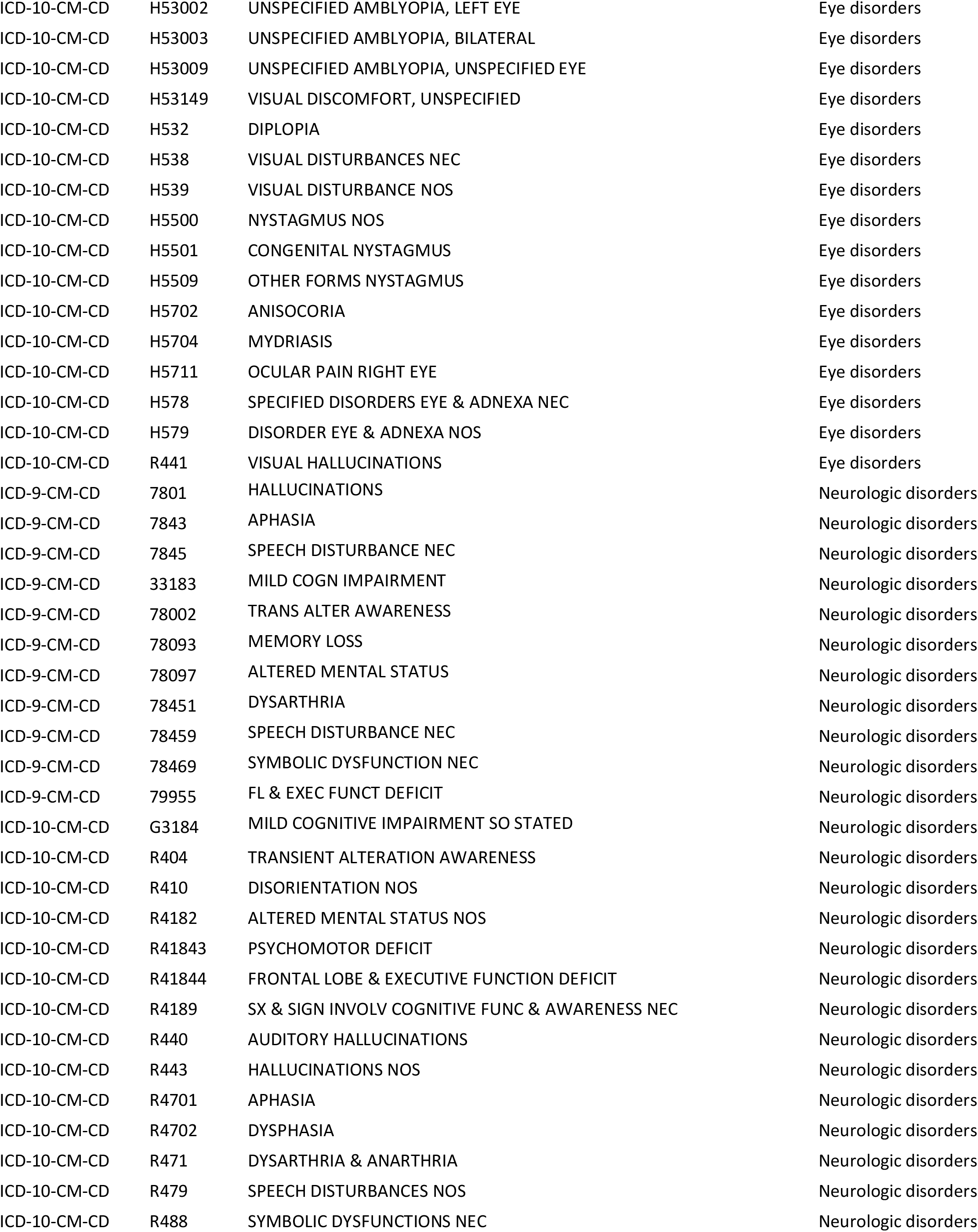
List of ICD10 codes used to determine presence of cALD.

